# Inconsistent Addiction Treatment for Patients Undergoing Cardiac Surgery for Injection Drug Use-Associated Infective endocarditis

**DOI:** 10.1101/19008482

**Authors:** Max Jordan Nguemeni Tiako, Seong Hong, Syed Usman Bin Mahmood, Makoto Mori, Abeel Mangi, James Yun, Manisha Juthani-Mehta, Arnar Geirsson

## Abstract

**Introduction:** Cases of injection drug use-related infective endocarditis (IDU-IE) requiring surgery are rising in the setting of the current U.S. opioid epidemic. We thus aimed to determine the nature of addiction interventions in the perioperative period.

**Methods:** This is a retrospective review of surgical IDU-IE from 2011 to 2016 at a tertiary care center in New Haven, Connecticut. The data collected included substances consumed recreationally, consultations by social work (SW), psychiatry, pharmacotherapy for addiction, and evidence of enrollment in a drug rehabilitation program upon discharge.

Among patients with active drug use (ADU), we compared the 24-month survival of those who received at least one form of addiction intervention to that of those who did not.

**Results:** Forty-two patients (75%) had active drug use. Among them, 22 used heroin. Forty-one patients (73.2%) saw SW, 17 (30.4%) saw psychiatry; 14 (25%) saw neither SW nor psychiatry.

Twenty-one patients (37.5%) received methadone, 6 (10.7%) received buprenorphine, 1 (0.02%) received naltrexone; 26 (46.4%) did not receive any pharmacotherapy. Fifteen patients (26.8%) attended a drug rehabilitation program, 13 (86.7%) of whom had seen SW and 8 (53%) psychiatry. Among patients with ADU, there was no statistically significant difference in survival between those who received at least one intervention and those who did not (p=0.1 by log rank).

**Conclusion:** Addiction interventions are deployed inconsistently for patients with surgical IDU-IE. Untreated substance use disorder and recurrent endocarditis are the leading cause of death in this population. Studying best-practices for perioperative interventions in IDU-IE and establishing protocols are of the upmost importance.

## Introduction

Heroin use has sharply increased in the United States.^1^ With the current opioid epidemic, data shows that individuals with prescription opioid use are 40 times more likely to inject heroin.^2,3^ Concurrently, the proportion of injection drug use-related infective endocarditis (IDU- IE) cases requiring valve surgery has increased significantly.^4,5,6^. Although surgical interventions play a major role in addressing IE refractory to pharmacotherapy,^7^ studies suggest that patients who use injection drugs (PUID) and undergo surgery for IDU-IE die mostly of causes related to untreated substance use disorder (recurrent endocarditis or drug overdose).^8,9^ In this study, we sought to describe the patterns of addiction treatment in PUID with IDU-IE requiring surgery.

## Methods

### Patient Population

We conducted a retrospective review of 56 consecutive patients who underwent cardiac surgery for IDU-IE from 2011 to 2016 at Yale New Haven Hospital, a tertiary care hospital in the United States. The patients were defined as having IE based on presentation with valvular disease of infectious etiology, as defined by the Center for Disease Control and Prevention (CDC), and adopted by the Society of Thoracic Surgeons and the Duke Criteria for infective endocarditis.^10^ Intravenous drug use was defined as a patient having a history of illicit drug use. Further chart review was performed to characterize the drug use as either ongoing or remote, as well as determining the types of drugs, based on patient reporting and collateral information from family members available in the medical record. The study was approved by the Yale Institutional Review Board and individual patient consent was waived.

### Data Sources

Data were collected through the local database extracted from the EMR and additional review of individual patients’ electronic medical record. Addiction treatment was defined as the aggregate of psychiatry and social work consults during inpatient stay and inpatient medication (buprenorphine, methadone, naltrexone, naloxone). Rehabilitation attendance upon discharge was confirmed via electronic health record review, specifically by searching discharge summaries, nursing notes and follow-up visit notes for mentions of rehabilitation attendance.

### Survival

24-month survival data was through linkage with the Connecticut state vital statistics data. Kaplan Meier curve characterized mid-term survival. A p value <0.05 was used to define statistically significant difference.

## Results

### Patient Characteristics

This study included 56 PUID who underwent surgical intervention for IDU-IE. The majority of patients were male (49, 87.5%) with average age of 44 ± 13 years. Prevalence of liver disease (23, 41.1%) was high, 22 patients (39.3%) had a history of prior stroke, and 15 (26.8%) had prior cardiac surgery, the majority of which had prior valve surgery (13/15, 86.7%). Seventeen patients (30.4%) received double-valve surgery, and 3 (5.4%) received triple-valve surgery (table 1).

**Table 1:** Pertinent patient characteristics

| Patient characteristics | All patients (total 56) | Patients with active drug use (total 42) |
| --- | --- | --- |
| Age | 44.1±13 | 42.6±13.8 |
| Caucasian race | 43 (76.8%) | 29 (69%) |
| Female sex | 7 (12.5%) | 4 (9.5%) |
| History of diabetes | 4 (7.4%) | 1 (2.4%) |
| History of hypertension | 19 (33.9%) | 11 (26.2%) |
| History of dialysis | 3 (5.4%) | 3 (7.1%) |
| History of stroke | 22 (39.3%) | 19 (45.2%) |
| Non-elective cases | 42 (75%) | 38 (90.5%) |
| History of cardiac surgery | 15 (26.8%) | 9 (21.4%) |

### History of Drug Use

In this population, 42 patients had active drug use at the time of their surgery, while the rest were noted to have a remote history of drug use. Among the 42, more than half of patients used heroin (22, 52.4%). Among the 22 patients who used heroin, over a third used heroin and cocaine only (8, 36.4%), and nearly a quarter (5, 22.7%) used heroin along with 2 or more other drugs (benzodiazepine, PCP, street suboxone, cocaine, Percocet, marijuana). Additionally, more than half of patients in the entire group of 56 had a concurrent history of tobacco use (29, 51.8%).

### Addiction Treatment Pattern

#### Specialty Consults

Among the 56 patients, 40 (71.4%) patients were seen by a social worker, and 17 (30.4%) were seen by a psychiatrist during their hospitalization. Fourteen patients were not seen by either a social worker or a psychiatrist; therefore, 25% of patients had no psychosocial consultation before being discharged (table 2).

**Table 2:** Summary of addiction interventions received by patients undergoing surgery for IE with active drug use or a remote history of drug use

| Addiction interventions | All patients (total 56) | Patients with active drug use (total 42) |
| --- | --- | --- |
| Methadone | 21 (37.5%) | 14 (33.3%) |
| Buprenorphine | 6 (10.7%) | 5 (11.8%) |
| Naltrexone | 1 (1.8%) | 1 (2.3%) |
| Naloxone | 14 (25%) | 13 (31%) |
| Social Work consults | 40 (71.4%) | 33 (78.6%) |
| Psychiatry consults | 17 (30.4%) | 15 (35.7%) |
| Rehabilitation attendance upon discharge | 15 (26.8%) | 10 (23.8%) |

Among the 42 patients who had active drug use, 33 (79%) were seen by a social worker, 15 (36%) were seen by a psychiatrist, and 8 were not seen by either a social worker or a psychiatrist; therefore, 19% of patients with active drug use had no psychosocial consultation before being discharged (table 2).

#### Pharmacotherapy

Pharmacotherapy was defined as the administration of methadone, buprenorphine, naltrexone or naloxone during the patient’s hospitalization. 21 patients (37.5%) were prescribed methadone, 6 patients (10.7%) were prescribed buprenorphine, 14 (25%) were prescribed naloxone, and 1 (0.02%) was prescribed naltrexone. 26 patients (46.4%) were not prescribed any of the aforementioned medications.

Among the 42 patients with active drug use, 14 (33%) patients received methadone, 5 (11.9%) received buprenorphine (including 2 who received both buprenorphine and methadone) and 1 (2.4%) received naltrexone. 18 (42.9%) patients total received at least 1 form of pharmacotherapy.

A total of 6 (14.3%) patients out of the 42 patients with active drug use received both pharmacotherapy and psychosocial interventions (table 2).

### Drug Rehabilitation Program Attendance

Fifteen patients were discharged to a drug rehabilitation facility. Thirteen (86.7%) patients discharged to a rehabilitation facility were seen by a social worker, and 8/15 (53.3%) patients discharged to a rehabilitation facility had been seen by a psychiatrist.

Among patients with active drug use, 10 patients were discharged to a drug rehabilitation facility. 9 (90%) were seen by a social worker and 7 (70%) were seen by a psychiatrist.

Conversely, among patients who did not go to a rehabilitation facility, 27 (65.9%) were seen by a social worker, and 9 (22%) were seen by a psychiatrist. The rehabilitation programs included 12-step group rehabilitation, narcotics anonymous, and adherence to daily methadone programs (table 2).

### Survival

Unadjusted mid-term survival showed no statistically significant (p=0.1 by log rank) difference between patients who received no treatment and those who received at least 1 form of intervention, including pharmacotherapy, psychiatry and social work consultations (Figure 1).

**Figure 1:**
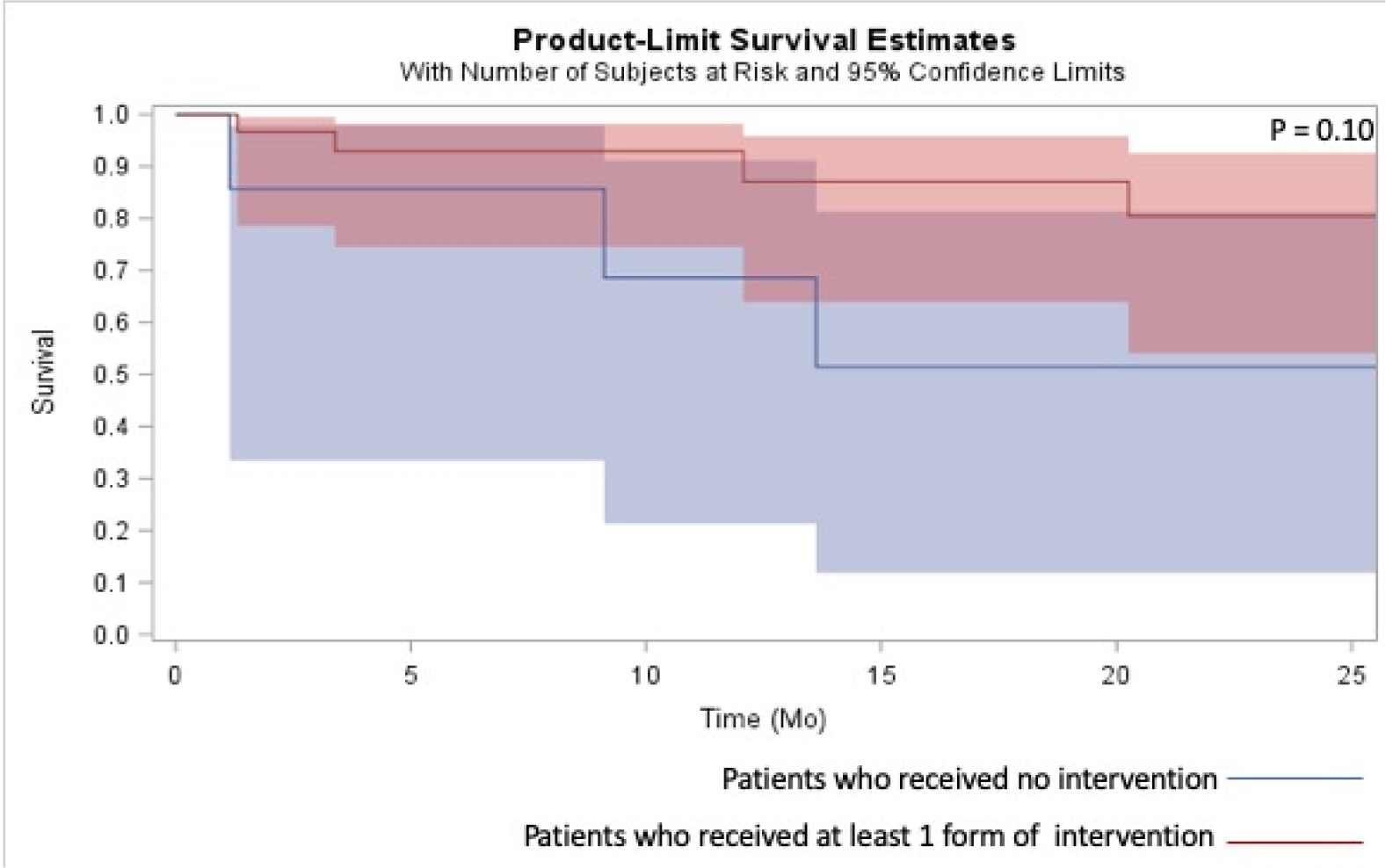
Kaplan-Meir curve of mid-term mortality comparing patients who received at least 1 addiction intervention and patients who did not receive any intervention

## Discussion

The aim of this study was to characterize patterns of addiction treatment at a tertiary center for PUID with IDU-IE requiring surgical intervention and their participation in drug rehabilitation programs upon discharge. We observed that there was a heterogeneous pattern of care provided given the lack of standardized protocols for addiction care. Currently, there are no guidelines for pharmacotherapy or usage of specialty consultations for addiction and psychosocial concerns surrounding the surgical management of IDU-IE.

We observed that psychiatrists were consulted as a post-operative care pathway. Methadone was most frequently prescribed as pharmacotherapy for addiction treatment postoperatively; however, harm reduction interventions through naloxone prescription were not deployed for all patients. As previously elaborated in a case-series for IDU-IE among PUID, their hospitalization is an important but often missed window of opportunity for medication-assisted therapy initiation.^11^ These findings are in line with prior studies suggesting that PUID hospitalized for IDU-IE receive suboptimal addiction treatment.^12^

An important finding in our study is the underutilization of transition to drug rehabilitation facilities. Notably, among 42 patients who had been actively using drugs, only 10 patients were discharged to a rehabilitation program. Most patients who were discharged to a rehabilitation program/facility were seen by a social worker and by a psychiatrist during their hospitalization, suggesting the importance of the role played by social workers and psychiatrists in the care of these patients.

As rates of IDU-IE requiring surgical intervention have been increasing throughout the opioid epidemic nationally,^5,13^ cardiac surgeons have been deliberating on the best way to care for these patients. Due to the risk of recurrence/reinfection associated with drug use^14^ and the limited nature of resources, different strategies have been proposed. A recent study highlighting right-sided endocarditis cases requiring surgery recommends staging operations, starting with a total tricuspid valvectomy, and a subsequent prosthetic valve implantation provided that patients are able to demonstrate abstinence from using drugs during a latency period of 6 months in between the two surgeries.^5^ This period of abstinence, however, is a state of vulnerability for patients, putting them at high risk of overdose should they relapse.^15^

Given that surgery for prosthetic valve endocarditis has been shown to have comparable outcomes to surgery for native valve endocarditis,^16^ some surgeons are inclined to opt for a comprehensive approach with immediate valve replacement.^17^ Bioethicists, however, raise the concern of resource limitation.^18^ In that vein, some suggest that contracts between patients and surgeons, akin to contracts between pain specialists and patients, may contribute to curbing return to drug use, and thus reinfection rates.^19^

However, this is a controversial position, and many oppose the notion of contracts dictating future access to life saving measures such as cardiac surgery, especially for such a marginalized population, given that addiction is the root problem.^20^ As our study demonstrates, peri-operative addiction management is not consistently employed in a large proportion of PUID. Several studies have shown presumed continued drug use to be the leading cause of death among PUID who underwent cardiac surgery for IDU-IE.^8^

Continued drug use is a consequence of addiction, and inherently increases the risk of reinfection. Addiction is a chronic neurobiological disorder, characterized in part by craving and withdrawal symptoms. Studies show that initiation of pharmacotherapy for treatment of opioid dependence in emergency room settings yields reduced self-reported illicit opioid use and decreased use of inpatient addiction treatment services.^21–23^ Additionally, studies suggest that for comprehensive addiction care for patients with opioid use disorder during hospitalization improves not only patient outcomes, but also provider outcomes.^24^ Other interventions such as long term residential addiction treatment and patient navigator programs have been shown to improve outcomes and reduce readmission rates for at risk patients, hence the importance of post-operative discharge to drug rehabilitation facilities or enrollment in rehabilitation programs.^25^ In cases where such resources may not be deployed, adequate outpatient follow-up after initiation of pharmacotherapy could contribute to sustaining improvement in outcomes.

### Limitations

This retrospective study was conducted at a single-center by observational electronic medical record chart review. Data on reoperation and recurrent endocarditis was not completely captured. For patients with a history of remote drug use but no indication of active use, the type of drugs consumed was not ascertained. Given that this is an observational study with a relatively small sample, the direct impact of medication initiation for addiction treatment and psychosocial consultations on rehabilitation attendance after discharge were not determined with statistical significance.

### Conclusion

Perioperative intervention for opioid use disorder in patients undergoing cardiac surgery for IDU-IE does not happen in a standardized fashion. Some patients receive pharmacotherapy and are seen by specialists equipped to address psychosocial factors, but not all. Given that mortality in this patient population is predominantly due to untreated substance use disorder, the concomitance of addiction treatment and surgical intervention through multidisciplinary care is of extreme importance. In the context of an opioid epidemic in the U.S., our findings argue for the development of a standard peri-operative addiction care pathway for this patient population.

## Data Availability

The data related to this manuscript will be made available upon request.

## Acknowledgments

### Sources of Funding

This study was funded by the Horace Stansel Jr Research Fund.

### Disclosures

Authors report no conflict of interests.

## Notes

### Competing Interest Statement

The authors have declared no competing interest.

### Author Declarations

All relevant ethical guidelines have been followed and any necessary IRB and/or ethics committee approvals have been obtained.

Any clinical trials involved have been registered with an ICMJE-approved registry such as ClinicalTrials.gov and the trial ID is included in the manuscript.

